# Using symptom-based case predictions to identify host genetic factors that contribute to COVID-19 susceptibility

**DOI:** 10.1101/2020.08.21.20177246

**Authors:** Irene V. van Blokland, Pauline Lanting, Anil P.S. Ori, Judith M. Vonk, Robert C.A. Warmerdam, Johanna C. Herkert, Floranne Boulogne, Annique Claringbould, Esteban A. Lopera-Maya, Meike Bartels, Jouke-Jan Hottenga, Andrea Ganna, Juha Karjalainen, Lifelines COVID-19 cohort study, The COVID-19 Host Genetics Initiative, Caroline Hayward, Chloe Fawns-Ritchie, Archie Campbell, David Porteous, Elizabeth T. Cirulli, Kelly M. Schiabor Barrett, Stephen Riffle, Alexandre Bolze, Simon White, Francisco Tanudjaja, Xueqing Wang, Jimmy M. Ramirez, Yan Wei Lim, James T. Lu, Nicole L. Washington, Eco J.C. de Geus, Patrick Deelen, H. Marike Boezen, Lude H. Franke

**Affiliations:** University of Groningen, University Medical Center Groningen, Department of Genetics, Groningen, The Netherlands; University of Groningen, University Medical Center Groningen, Department of Cardiology, Groningen, The Netherlands; University of Groningen, University Medical Center Groningen, Department of Psychiatry, Groningen, The Netherlands; University of Groningen, University Medical Center Groningen, Department of Epidemiology, Groningen, The Netherlands; Structural Computational Biology unit, EMBL, Heidelberg, Germany; Department of Biological Psychology, FGB, Vrije Universiteit Amsterdam, Amsterdam, The Netherlands; Amsterdam Public Health research institute, Amsterdam UMC, Amsterdam, The Netherlands; Institute for Molecular Medicine Finland, University of Helsinki, Helsinki, Finland; Broad Institute of MIT and Harvard, Cambridge, MA, USA; Analytic and Translational Genetics Unit (ATGU), Massachusetts General Hospital, Boston, MA, USA; MRC Human Genetics Unit, Institute of Genetics and Molecular Medicine, University of Edinburgh, Edinburgh, UK; Department of Psychology, University of Edinburgh, 7 George Square, Edinburgh EH8 9JZ; Medical Genetics Section, Centre for Genomic and Experimental Medicine, Institute of Genetics and Molecular Medicine, University of Edinburgh, Edinburgh, UK; Helix, 101 S Ellsworth Ave Suite 350, San Mateo, California 94401, USA; Department of Genetics, University Medical Centre Utrecht, P.O. Box 85500, 3508 GA, Utrecht, The Netherlands

**Author notes:** shared first authors, these authors contributed equally to this work (Irene V. van Blokland, Pauline Lanting, Anil P.S. Ori, Judith M. Vonk, Robert C.A. Warmerdam). shared last authors (H. Marike Boezen, Lude H. Franke). **Corresponding author**: Prof. Lude Franke, Department of Genetics, University Medical Center Groningen, Groningen, The Netherlands.

**Keywords:** COVID-19, disease prediction, SARS-CoV-2, self-reported symptoms, GWAS, host genetic factors

## Abstract

Epidemiological and genetic studies on COVID-19 are currently hindered by inconsistent and limited testing policies to confirm SARS-CoV-2 infection. Recently, it was shown that it is possible to predict potential COVID-19 cases using cross-sectional self-reported disease-related symptoms. Using a previously reported COVID-19 prediction model, we show that it is possible to conduct a GWAS on predicted COVID-19, and this GWAS benefits from the larger sample size to provide new insights into the genetic susceptibility of the disease. Furthermore, we find suggestive evidence that genetic variants for other viral infectious diseases do not overlap with COVID-19 susceptibility and that severity of COVID-19 may have a different genetic architecture compared to COVID-19 susceptibility. Our findings demonstrate the added value of using self-reported symptom assessments to quickly monitor novel endemic viral outbreaks in a scenario of limited testing. Should there be another outbreak of a novel infectious disease, we recommend repeatedly collecting data of disease-related symptoms.

## Introduction

The Coronavirus Disease 2019 (COVID-19) caused by Severe Acute Respiratory Syndrome Coronavirus-2 (SARS-CoV-2) has rapidly spread across the globe, posing a large burden on individuals, healthcare systems, and societies as a whole. At the time of writing, more than 16 million infections and 650,000 deaths have been reported worldwide.^1^ The symptoms and disease severity of COVID-19 vary^2^, ranging from asymptomatic or nonspecific symptoms to severe illness with hospital admission and death. While the scientific community is rapidly gaining more understanding of the pathophysiology of COVID-19^3,4^, many questions remain about the etiology of the disease and what factors are driving the interindividual variability in pathophysiology.

It is known that individual genetic differences in the human host contribute to immune function and response to common infectious agents.^5,6^ Genome-wide association studies (GWAS) have, for example, identified susceptibility loci for multiple common infections.^7^ The identification of genetic factors can lead to new insights into disease mechanisms and help improve vaccination strategies by optimizing vaccine-induced protection. For this reason, the COVID-19 host genetics consortium (C19HG) was established to discover and study the human genetic variants that modulate the susceptibility of developing COVID-19 symptoms and COVID-19 severity.^8^ However, the magnitude of the pandemic, limited testing capacity and inconsistent testing policies have likely resulted in an underrepresentation of the number of true cases. Using only confirmed cases reduces the power of any GWAS to detect associations and may be a source of bias.

Recently, a model was published that predicts the potential presence of COVID-19 based on self-reported disease-related symptoms, which we will refer to as the Menni COVID-19 prediction model.^9^ We investigated if potential COVID-19 predicted based on symptoms can help accelerate the search for host genetic factors that contribute to the susceptibility of developing COVID-19 symptoms, which we will refer to as COVID-19 susceptibility, and the heterogeneity of COVID-19 severity. First, we confirmed that the Menni COVID-19 model can identify cases with laboratory confirmed SARS-CoV-2 infection in three independent cohorts. As existing COVID-19 prediction models used features not available in our cohorts, we generated a COVID-19 prediction model optimized to the phenotypes described in our cohorts. Second, as part of the C19HG consortium, we performed genetic analyses on predicted COVID-19 (1,865 cases and 29,174 controls, **Figure 1**) to search for host genetic factors that contribute to COVID-19 susceptibility and explored possible downstream effects of the loci identified. To assess the validity of the predicted COVID-19 phenotype, we compared these results to the GWAS meta-analyses results based on confirmed COVID-19. We also compared our findings to previously reported genetic associations with several viral infectious diseases to look for genetic factors shared between COVID-19 susceptibility and other viral infectious diseases.

**Figure 1.**
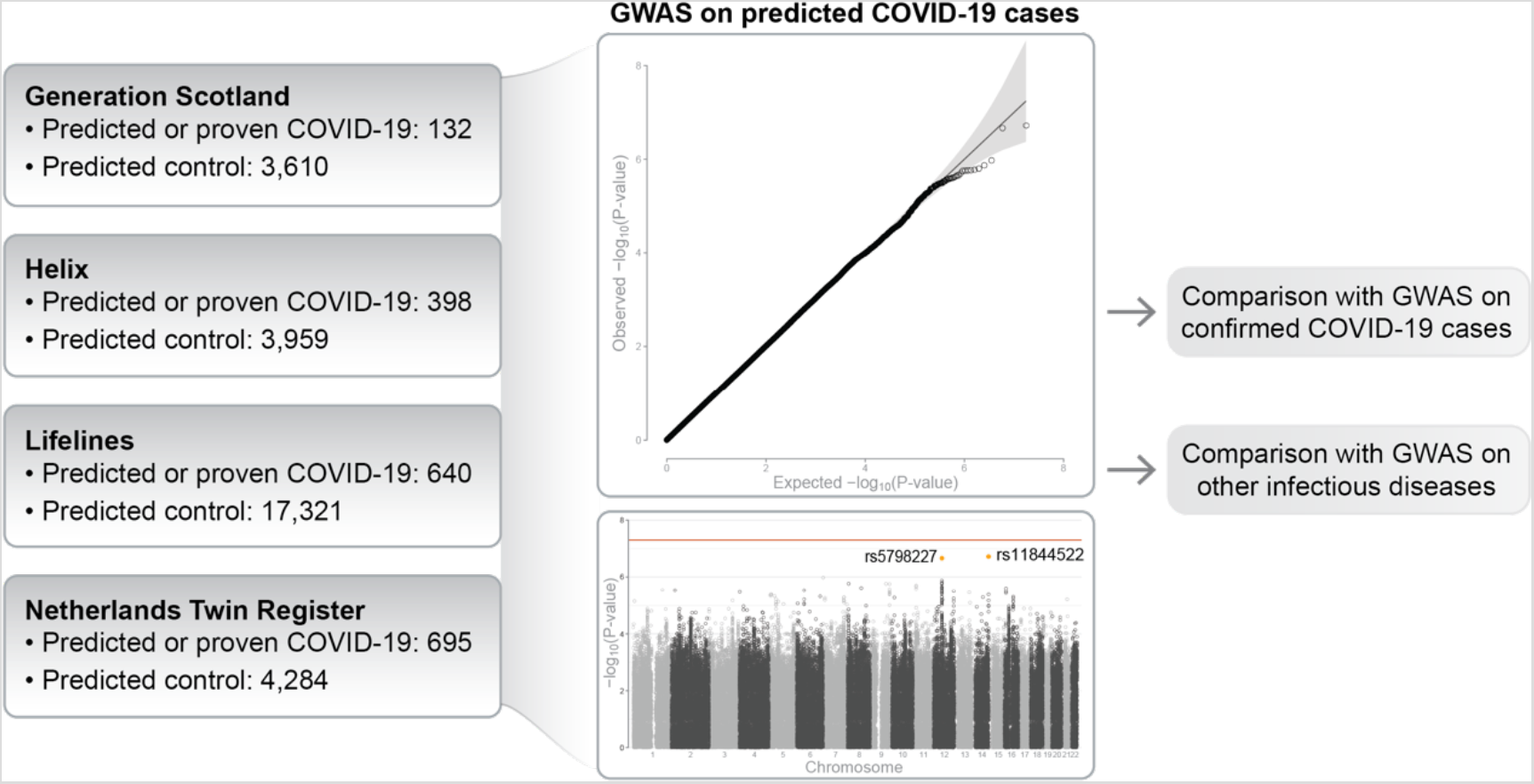
Overview of the main analysis

## Materials and Methods

### Data collection and preparation

The Generation Scotland cohort consists of individuals over the age of 18 from the Generation Scotland biobank. The CovidLife survey was initially launched on April 17, 2020 with a few hundred individuals to make sure the survey process was working well. The following week, the survey was rolled out by email and letter to all of the current Generation Scotland volunteers. Volunteers were asked questions about the impact the pandemic had on their life and included questions on education, mental health, wellbeing and more.

The Helix cohort consists of individuals from the Helix DNA Discovery Project, an unselected population of adults from across the United States^13^. COVID-19 questionnaires were emailed to participants in April and May of 2020. The questionnaire format was based on example surveys and suggested symptoms and pertinent information compiled by the C19HG^14^.

The Lifelines COVID-19 cohort consists of individuals from the Lifelines population cohort and the Lifelines NEXT birth cohort in the Northern part of the Netherlands^5^ ^6^. Within the Lifelines COVID-19 cohort, questionnaires were sent out to participants over the age of 18 years via email on a weekly basis starting March 30, 2020. Items about COVID-19 infection and perceived symptoms, drug use, mental health and vaccination status were questioned weekly. A comprehensive cohort description has been described previously^15^.

The Netherlands Twin Register (NTR) consists of members of twin families that had been registered as willing to participate in survey, biobank and experimental research. NTR participants aged 16 years or older (range 16-95) received an online questionnaire at the end of April (wave 1) or the middle of May (wave 2). The questionnaire was modelled on the Lifelines survey and contained items about COVID-19 testing, diagnosis and treatment of COVID-19, perceived flu-like symptoms, drug use, past and present chronic diseases, household composition, work setting and the impact of the corona crisis on their mental health and lifestyle behaviours.

### Application of the Menni COVID-19 prediction model

To replicate and apply the Menni COVID-19 prediction model, symptoms described using non-binary answers (5- or 7-point Likert scale answers) were recoded into binary responses using arbitrary cut-off values. An overview of all available symptoms, corresponding answer scales and the cut-off values are shown in **Table S1**. The symptom ‘skipped meals’ was not inquired about in all of the cohorts. This symptom could not be substituted by another symptom in Lifelines. It was substituted by ‘severe loss of appetite’ in NTR, by ‘decreased appetite’ in Helix and by ‘lack of appetite’ in Generation Scotland. ‘Severe or significant persistent cough’ was substituted with the maximum response values of ‘cough without sputum’ and ‘cough with sputum’ in Lifelines, by ‘dry cough lasting at least 20 days’ in Helix, ‘coughing – any’ with an alternative high cut-off in NTR and ‘dry cough’ in Generation Scotland. ‘Severe fatigue’ was substituted with the minimum response values of the following fatigue items in Lifelines: ‘feeling tired’, ‘feeling tired quickly’ and ‘feeling physically exhausted’. In NTR ‘feeling tired’ was used with an alternative higher cut-off to recode the 5-point scale into binary. Generation Scotland substituted ‘severe fatigue’ with ‘fatigue/tiredness’.

For replication, we applied the Menni COVID-19 prediction model (i.e. predicted COVID-19 score = −1.32 – (0.01 * age) + (0.44 * male sex) + (1.75 * loss of smell and taste) + (0.31 * severe or significant persistent cough) + (0.49 * severe fatigue) + (0.39 * skipped meals)) to the Helix, Lifelines and NTR cohorts separately. The different cut-offs that were applied by the cohorts when applying the Menni model to the datasets prior to running the GWAS are shown in **Table S2** We calculated the predicted probability of COVID-19 according to exp(predicted COVID-19 score)/(1+exp(predicted COVID-19 score)). The predictive properties were tested using an ROC analysis. Sensitivity, specificity, positive predictive value (PPV) and negative predictive value (NPV) were calculated based on a predicted probability higher than 0.50 to define a positive predicted case.

### Attempt to improve the Menni COVID-19 prediction model

The three cohorts with self-reported SARS-CoV-2 reverse-transcription PCR (RT-PCR) test outcomes available (Helix, Lifelines and NTR) were used in an attempt to improve the Menni COVID-19 prediction model. Self-reported symptoms that were present in all three cohorts were used. Symptoms were reported on a 5-point or 7-point Likert-scale (Lifelines COVID-19 and NTR cohorts) and binary scale (Helix cohort). To categorize all symptoms into a binary variable, we assessed the appropriate cut-off values in the Lifelines COVID-19 cohort for each self-reported symptom by performing a logistic regression on subjects with a positive test outcome (n = 56) compared to subjects with a negative test outcome (n = 586). In these models, each symptom was investigated separately by using two dummy variables indicating low and intermediate/high symptom severity with the reference being the absence of the symptom. If only intermediate/high symptom severity was significantly associated with a positive test, we used this value as cut-off. If both low and intermediate/high symptom severity were significant, we used low severity as cut-off.

The symptoms selected for this model had to be present for the entire data-collection period in all three cohorts or a proper substitution had to have been possible, resulting in the following symptoms being selected: coughing-any, diarrhea/stomach ache, difficulty breathing, fever, loss of smell/taste, runny nose, sore throat and tired-any. Subsequently, we performed forward and backward stepwise logistic regression in the Lifelines COVID-19 Cohort to construct the model most predictive for a positive test outcome (p-in = 0.10 and p-out = 0.10). The predictive properties were tested using an ROC analysis. Sensitivity, specificity, PPV and NPV were calculated based on the predicted probability, favouring an optimal PPV. We then validated this model in the Helix and NTR cohorts.

### Genome-wide association analysis

We performed GWAS for predicted COVID-19 case-control status as part of the COVID-19 Host Genetics Initiative (C19HG) with a total of 1865 cases and 29174 controls (https://www.covid19hg.org/results/). All cohorts consist of individuals of European ancestry. See **Table S3** for the full phenotype description and detailed analysis plan. Additional details on cohort level GWAS and C19HG meta-analysis are provided in the **Supplementary Methods**.

### Enrichment analysis

We selected all variants with a p-value ≤ 5×10^−4^ and used DEPICT^10^ with default settings to search for enrichment in pathways and protein-protein interactions. We used a false discovery rate of 0.05.

### Processing of GWAS results

We downloaded the results of meta-analyses from the C19HG website (https://www.covid19hg.org/results/) (genome assembly GRCh37, retrieved on 02-07-2020) for *B2*: Hospitalized COVID-19 vs. population, *C1*: COVID-19 vs. self-reported negative, *C2*: COVID-19 vs. population and *D1*: Predicted COVID-19 from self-reported symptoms vs. predicted or self-reported non-COVID-19. Hereafter, we added RSIDs where both the genomic location and alleles matched to a variant from dbSNP https://ftp.ncbi.nih.gov/snp/redesign/latest_release/VCF/GCF_000001405.25.gz, retrieved on June 30th). Variants in the *D1* meta-analysis were filtered on MAF>0.01 (all_meta_AF column), after which we performed p-value informed LD pruning, also called clumping, using PLINK (v1.90b6.10 64-bit, –-clump) and the European population from the 1000 Genomes Project (phase 3) as a reference panel. For clumping, thresholds on the linkage disequilibrium (--clump-r2) and genomic distance (--clump-kb) were set to an R^2^ of 0.2 and a distance of 250 kb respectively. In GWASs other than the *D1* analysis, the maximum p-value of index variants (--clump-p1) was set to 510-8. All other parameters were left as their default values.

### Comparison between predicted COVID-19 and three other GWASs

For the *D1* GWAS, the top 20 independent SNPs were selected. The same SNPs were selected from the *C1, C2* and *B2* GWASs to determine if their effects replicated. The same was done using the independent genome-wide significant hits from the *C1, C2* and *B2* analyses. The results from these queries were aggregated and summarized.

### Comparison of COVID-19 GWASs with previously reported associations in viral infection phenotypes

First, genome-wide significant variants (P ≤ 5×10^−8^) were selected for common viral infection phenotypes from the NHGRI-EBI GWAS Catalog (accessed July 7, 2020)^22^. Next, the individual SNPs corresponding to each association were queried from the *D1, C1, C2* and *B2* COVID-19 GWASs. For every viral infection SNP that we found in one of the four COVID-19 GWASs, we determined if the SNP replicated, dictated by the p-value of the association in the respective COVID-19 GWAS, the Bonferroni-corrected significance level calculated from the number of SNPs for a viral infection trait and an a priori Bonferroni adjusted alpha of 0.05.

To get a more concrete indication whether or not the COVID-19 GWASs showed an increased signal of previously reported viral infection associations, quantile-quantile plots were made and accompanying genomic inflation factors (λ) were calculated in the selection of SNPs that have previously been reported to be associated with the various viral infection traits. A significance value for every λ was calculated by simulating 1000 expected λ-values, calculating the consequent Z-score for the observed λ, and determining a two-tailed p-value. λ-values were simulated by sampling n values from a χ^2^-distribution (k = 1), where n corresponds to the number of p-values used to calculate the observed λ-value.

## Results

### Description of cohorts

The Generation Scotland, Helix, Lifelines and Netherlands Twin Register (NTR) cohorts include a total of 168 (0, 27, 56 and 85, respectively) positively tested COVID-19 cases and 1157 (0, 189, 586 and 382, respectively) negatively tested controls. Descriptive statistics of the cohorts are provided in **Table S4**

### Replication of the Menni COVID-19 prediction model in Helix, Lifelines and NTR

We replicate the Menni COVID-19 prediction model as it was published, i.e. including age and gender and with a predicted probability higher than 0.50 to define a positive predicted case. **Table 1** presents **t**he model diagnostics of the replication of the Menni COVID-19 prediction model in the three independent cohorts. The Menni model yields an area under the curve (AUC) ranging between 0.79 and 0.86 across the three cohorts, similar to the performance reported in the original study.

**Table 1.** Model diagnostics of the Menni COVID-19 prediction model in Helix, Lifelines and NTR.

| Cohort | AUC (95% CI) | Sensitivity | Specificity | Positive predictive value | Negative predictive value |
| --- | --- | --- | --- | --- | --- |
| Helix | 0.79<br>(0.725-0.869) | 0.481 | 0.905 | 0.419 | 0.924 |
| Lifelines | 0.824<br>(0.758-0.890) | 0.446 | 0.951 | 0.463 | 0.947 |
| NTR | 0.864<br>(0.822-0.905) | 0.415 | 0.936 | 0.596 | 0.876 |

The model: $-1.32 - 0.01 \cdot \text{age} + 0.44 \cdot \text{male sex} + 1.75 \cdot \text{loss of smell or taste} + 0.31 \cdot \text{severe or significant persistent cough} + 0.49 \cdot \text{severe fatigue} + 0.39 \cdot \text{skipped meals}$ . A predicted probability cut-off of $> 0.50$ is used to define a positive predicted case.

### Phenotypic associations and prevalence of core predicted COVID-19 symptoms in Lifelines

In the Lifelines COVID-19 cohort, we observe that individuals who were tested for SARS-CoV-2 more often report working in essential occupations, for example in healthcare, and were generally younger. Individuals who tested positive for an infection report having been in contact with another infected individual (62.5%) more often than individuals with a negative test outcome (19.3%) (**Table S5**). Furthermore, infected individuals were already reporting symptoms of fever, loss of smell or taste, fatigue and coughing at higher frequencies even before testing positive and continued to report these symptoms thereafter (**Figure 2**). Individuals with a negative test outcome reported few to no symptoms of fever and loss of smell or taste, overall. Individuals predicted for potential COVID-19 show a pattern of reported symptoms more similar to that of individuals with a positive test outcome. While, as expected, the prevalence of loss of smell or taste, fatigue and coughing is highest at the time of the positive test, these symptoms are also still reported afterwards. For predicted COVID-19 cases, fewer symptoms of fever are reported than for cases with a positive test outcome.

**Figure 2.**
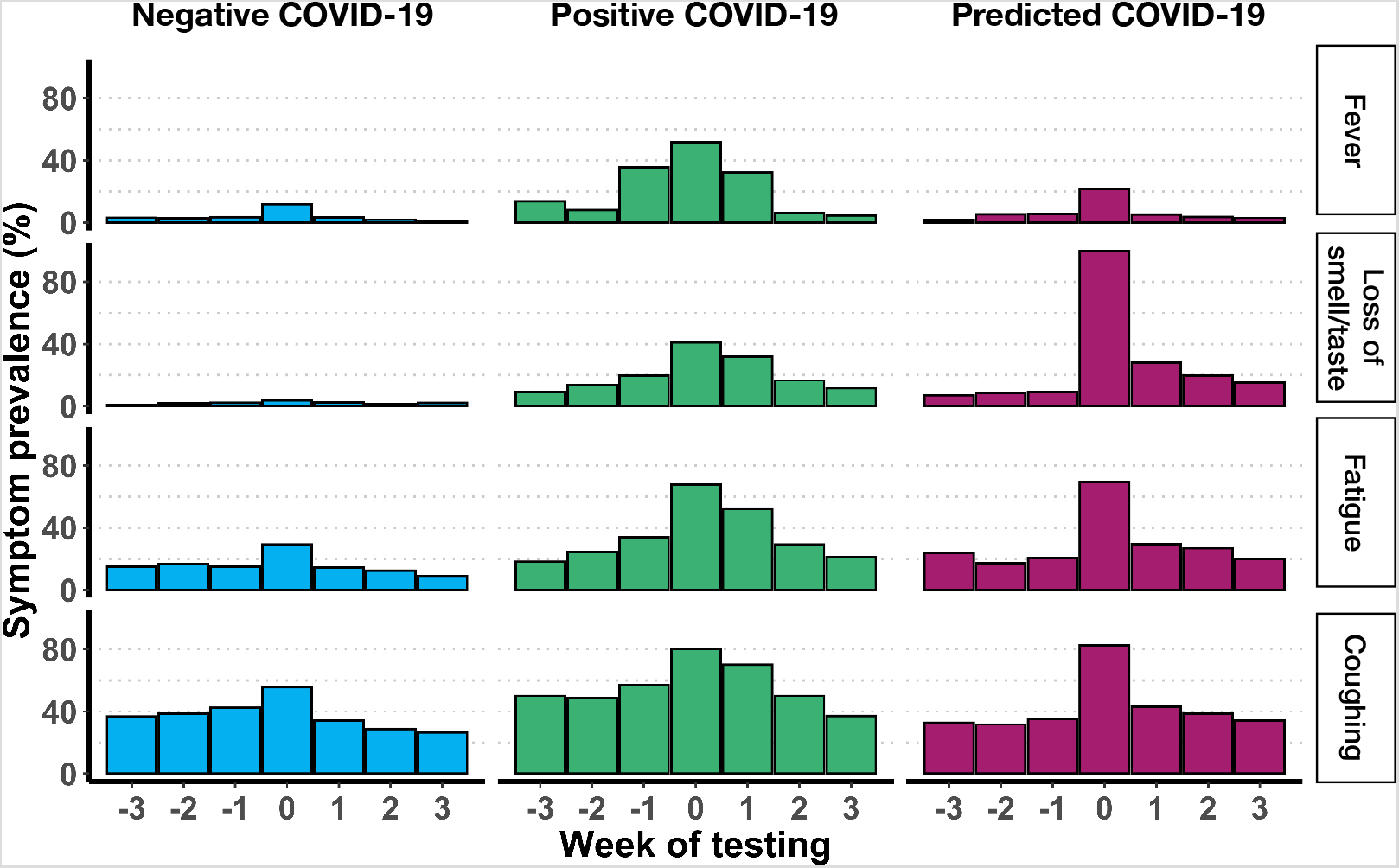
Prevalence of self-reported symptoms before and after testing negative, testing positive and predicted COVID-19 in the Lifelines cohort. To compare self-reported symptoms between cases with a negative test outcome (blue), a positive test outcome (green) and a predicted case status (purple), we aligned weekly reported symptoms for each individual to the week of testing (i.e. week 0). For each week, symptom prevalence (y-axis) was calculated and visualized in relation to the week of testing (x-axis). Shown are symptoms used as input in the Menni COVID-19 prediction model and fever.

To gain insights into disease associations within the group of predicted COVID-19 cases, we explored if predicted cases report specific pre-existing conditions more often than the controls in our GWAS. Here we observe a positive association with self-reported lung disease, chronic muscle disease, psychiatric illness, cancer and neurological disease (**Figure S1**). This was not observed when comparing patients with a positive versus negative test outcome. While this may indicate that predicted COVID-19 cases present a different profile of pre-existing conditions as compared to true cases, the comparison between predicted and positive COVID-19 cases remains biased due to testing policies and the small number of cases with a positive test outcome.

### A new Lifelines prediction model for COVID-19 yields similar performance

Using self-reported symptoms of 56 positive and 586 negative test outcome cases in the Lifelines cohort, we next attempted to improve on the Menni COVID-19 prediction model. The cut-off values used are presented in **Table S6**. The best prediction model was: −4.497 + 1.032 ✕ cough + 2.042 ✕ fever + 2.145 ✕ loss of smell or taste. The diagnostics of this Lifelines model are presented in **Table S7a, S7b** and **S7c**. A comparison of the case/control-predictions stratified by positive/negative test of the Lifelines and Menni COVID-19 prediction models is presented in **Table S8**. Overall, the prediction accuracies of the two models are comparable. As the Menni COVID-19 prediction model was developed and validated in two larger cohorts, we decided to continue with case prediction based on the Menni COVID-19 prediction model in the subsequent GWAS.

### The first GWAS of predicted potential COVID-19

We conducted a GWAS meta-analysis on 1,865 predicted cases and 29,174 controls across four independent cohorts. The full summary statistics of our analysis are available for download online on the C19HG website.^8^ The results of the top 20 (P < 5.1×10^−6^) independent single nucleotide polymorphisms (SNPs) for predicted COVID-19 are shown in **Figure 3**. Suggestive evidence of association with predicted COVID-19 was found for two SNPs (rs11844522, p = 1.9×10^−7^; rs5798227, p = 2.2×10^−7^) (**Figure S2**). Downstream analysis using DEPICT^10^ ascertained that protein-protein interactions with the solute carrier family 25 member 6 gene (*SLC25A6)* are significantly enriched (p = 8.6×10^−6^). Full results are provided in **Table S9**.

A comparison of the top 20 SNPs for predicted COVID-19 (*D1: Predicted COVID-19 from self-reported symptoms vs. predicted or self-reported non-COVID-19*) with three other COVID-19 phenotypes showed three SNPs to be associated with the same direction of effect (rs13288295 in *C1: COVID-19 vs. self-reported negative*, rs75517918 in *C2: COVID-19 vs. population* and rs143825287 in both *C1* and *C2*) and one SNP to be associated with the opposite direction of effect (rs11844522 in *B2: Hospitalized COVID-19 vs. population*), based on a p-value threshold of 0.05 (**Figure 3**).

**Figure 3.**
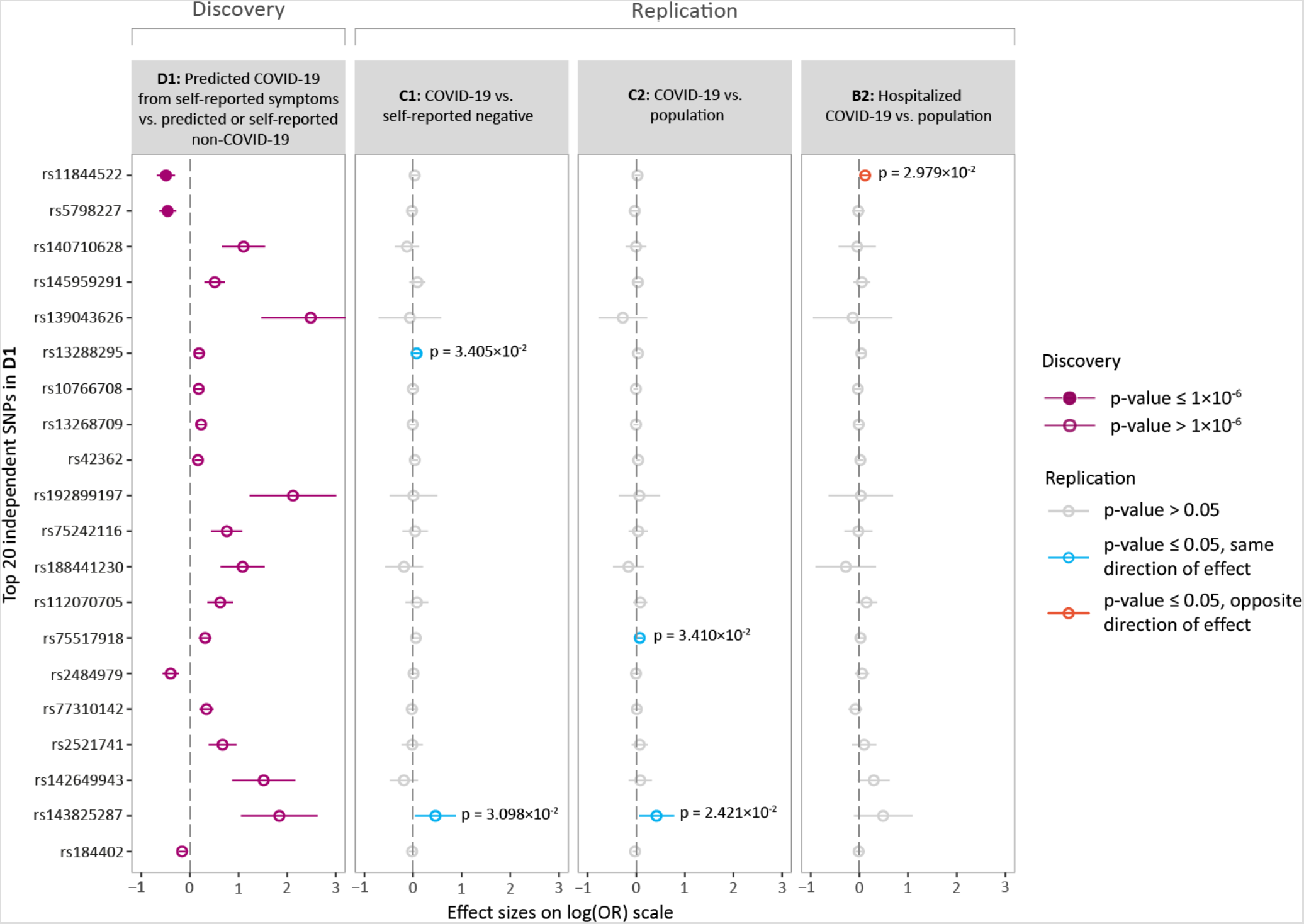
Overview of the top loci associated with predicted COVID-19. Shown are the effect size estimates of the top 20 independent SNPs associated with predicted COVID-19 (*D1)* and each of their associations with *C1* (COVID-19 vs. self-reported negative), *C2* (COVID-19 vs. population) and *B2* (Hospitalized COVID-19 vs. population). The effect sizes are shown with the risk allele odds ratio (OR) on a log-scale with a corresponding 95% confidence interval (CI). Colours indicate various p-value thresholds as described in the figure legend.

The meta-analyses of the *C2* GWAS showed an independent genome-wide significant association on locus 3p21.31 (rs35652899, p = 9.5×10^−11^). The *B2* GWAS also showed two approximately independent genome-wide significant associations, of which the most significant is in high linkage disequilibrium with the associated variant from the *C2* analysis (rs35044562, p = 3.1×10^−15^, R^2^ = 0.97). A comparison of these results to the *D1* GWAS showed that both these associations with closely linked variants did not replicate at a significance level of 0.05 (p-values 0.18 and 0.22, respectively).

Next, we examined whether previously reported genetic associations with common viral infections share any overlap with the variants identified by our GWAS on COVID-19 susceptibility. After querying the NHGRI-EBI GWAS Catalog, we further investigated 270 genome-wide significant SNPs associated with known viral infections. Here we observe no evidence of overlap with predicted COVID-19 at the level of individual SNPs (a priori Bonferroni-adjusted alpha = 0.05, **Figure S3**). Furthermore, there was no overall increase in genomic inflation (λ) when considering all 270 SNPs jointly for any of the four GWAS phenotypes (λ = 0.815, p = 0.2 *D1*, λ = 1.234, p = 0.1 *C1*, λ = 1.110, p = 0.5 *C2*, λ = 0.780, p = 0.1 *B2*, respectively) (**Figure S4**).

## Discussion

We investigated if symptom-based prediction of potential COVID-19 cases can aid in the search for host genetic factors that contribute to COVID-19 susceptibility. We confirm that self-reported disease-related symptoms are useful for prediction of infection status and report the first genome-wide association analysis on predicted potential COVID-19 in the C19HG consortium. We find suggestive evidence for rs11844522 and rs5798227 to be associated with predicted COVID-19, but observe no evidence for overlap with known genetic associations with common viral infections.

Individuals identified with the Menni COVID-19 prediction model and used in our GWAS could include cases that are not confirmed positive cases for COVID-19. While the Menni prediction model has good predictive properties, with AUCs ranging between 0.74 and 0.86 in the included cohorts, it yielded lower sensitivities (0.42 to 0.48) and positive predictive values(0.42 to 0.60). This indicates that a significant proportion of COVID-19 cases will be missed by the prediction model (false negatives) and that positive predicted cases will include false positives. As our attempt to improve this model was unsuccessful, this remains an avenue to explore for future work. Symptom prevalence before and after testing in Lifelines suggests that repeated self-report assessments of disease-related symptoms may offer finer resolution to further increase prediction accuracy.

The probable COVID-19 phenotype can help increase the number of cases for genetic analyses of COVID-19. While GWAS can benefit from larger sample sizes, caution should be taken when applying more loose phenotyping, as such an approach can produce a smaller and less-specific genetic signal.^11^ The predicted positive COVID-19 cases could include false positive cases who have underlying conditions, such as other viral infections, that share symptomatology with the symptoms included in the prediction model. This may have subsequently confounded our GWAS, yielding results that are less specific for COVID-19 and more related to genetic susceptibility to general immune defense or potentially even conditions un-related to COVID-19. Out of the 270 genome-wide significant hits for eight other infectious diseases, only five replicate, of which one in the *D1* phenotype had the lowest p-value of 3.077×10^−4^. Additionally, the calculated genomic inflation factors showed no inflation for viral infection SNPs across any of the four COVID-19 phenotypes. Based on these first results, we observe minimal overlap of our COVID-19 symptom SNPs with SNPs previously reported to be associated with viral infection phenotypes.

The outcomes of the GWAS meta-analysis of predicted potential COVID-19 showed suggestive association with the SNPs rs11844522 and rs5798227. Interestingly, rs11844522 is in a locus comprising immunoglobulins, and the closest mapping gene (*IGHV3-7*) to rs11844522 is part of a gene family that is enriched in total VDJ expression of COVID-19 patients in single-cell transcriptomic data^12^. rs11844522 replicated with an opposite direction of effect in the *B2* phenotype, and this is likely explained by the *B2* phenotype, which focused on COVID-19 severity (i.e. susceptibility to a poor outcome) rather than COVID-19 susceptibility (i.e. susceptibility to developing COVID-19 symptoms). The lack of genome-wide significant SNPs could largely be explained by our limited number of cases compared to the *C2* analyses (*D1*: 1,865 vs. *C2*: 6,696). Furthermore, among the 20 most-significant top-variants, three were identified and replicated (at p < 0.05) in *C1* or *C2*, all with the same direction of effect.

A comparison between COVID-19 GWASs showed that GWAS *D1* was unable to replicate the top genome-wide significant hit of the *C2* GWAS with positively tested cases. Interestingly, a closely linked variant in the *B2* analysis, which considers COVID-19 severity, is even more significant than the *C2* top variant (rs35044562: p = 3.1×10^−15^ and rs35652899: p = 8.6×10^−10^, respectively, R^2^ = 0.97). In the *C2* analysis, only the COVID19-Host(a)ge cohort, which contributed the largest number of cases, showed a genome-wide significant association with the top variant at this locus (p = 8.2×10^−11^), while other cohorts all showed much less significance (UK Biobank: p = 1.3×10^−3^; other cohorts: p > 0.01). Looking into this discrepancy revealed that the COVID19-Host(a)ge cohort focusses on severe COVID-19 patients, which is indicated by the fact that the cases contributed by this cohort in *C2* completely overlap with hospitalized cases contributed to the *B2* GWAS. A similar observation can be made for the UK Biobank, for which the number of hospitalized cases in *B2* constitute 66% of cases in *C2*, an observation that explains the increased significance for the association in this cohort compared to others. Taking these observations into account, it seems reasonable to assume that the reported variants on the 3p21.31 locus are more likely to be associated with COVID-19 severity than COVID-19 susceptibility. Therefore, no conclusion can be made on the performance of the predicted COVID-19 phenotype as a proxy for COVID-19 susceptibility based solely on the absence of an association with the 3p21.31 locus.

Downstream DEPICT analysis of the GWAS outcomes identified a significant enrichment of protein-protein interactions with *SLC25A6*. This gene encodes adenine nucleotide translocator 3 (ANT3), which is a core component of the mitochondrial permeability transition pore (MPTP) and is involved in apoptosis. *SLC25A6* is downregulated in human cytomegalovirus infection and associated with influenza virus–induced apoptosis.^10,11^ This indicates this gene might also be relevant to COVID-19 susceptibility.

### Limitations

There are multiple limitations of using predicted COVID-19 cases that we need to consider. Firstly, the training data might not be fully representative of the whole spectrum of COVID-19 symptoms since testing of putative cases in the early months of the pandemic was mostly restricted to patients with a more severe phenotype. Individuals with essential occupations, for example healthcare professionals, were also more frequently tested at the beginning of the pandemic. Secondly, some symptoms are also present in common chronic diseases, for example “loss of smell and taste” is frequent among patients with a neurological disorder. Indeed, a preliminary analysis of the Lifelines data showed enrichment of patients with preexisting conditions in the predicted COVID-19 cases as compared to controls but no enrichment in the confirmed COVID-19 cases compared to confirmed negative cases, indicating that these individuals might be incorrectly predicted as COVID-19 cases by the Menni COVID-19 prediction model based on their symptoms (**Figure S1**). Thirdly, the prevalence of COVID-19 might be different among different populations and cohorts. The false positive rates of the prediction models are likely to be larger if the prevalence of COVID-19 is small compared to other infectious diseases that often have similar symptoms.

### Conclusions

We show that it is possible to conduct a GWAS on predicted COVID-19. As GWAS of COVID-19 will benefit from larger samples, predicted COVID-19 can help increase the number of cases in GWASs, which will help to generate new insights into the genetic architecture of COVID-19 susceptibility. While the current predicted COVID-19 GWAS did not find genome-wide significant loci, our work does serve as a proof of concept. First analyses suggest that genetic variants involved in other viral infectious diseases do not overlap with COVID-19 susceptibility and that COVID-19 severity may have a partially different underlying genetic architecture. Our findings furthermore demonstrate the added value of using self-reported symptom assessments to quickly monitor the activity of novel endemic viral outbreaks in a scenario of limited testing. Should there be another outbreak of a novel infectious disease, we recommend collecting longitudinal data on disease-related symptoms.

## Data Availability

Non-identifiable information from the GS:SFHS cohort is available to researchers in the UK and to international collaborators through application to the GS Access Committee. GS operates a managed data access process including an online application form (http://www.gsaccess.org/) and proposals are reviewed by the GS Access Committee.
The Helix data were collected under IRB Protocol #20170748. Data are available from the corresponding author on reasonable request.
The Lifelines data analysed in this study was obtained from the Lifelines biobank, under project application number ov20_0554. Requests to access this dataset should be directed to Lifelines Research Office.
The NTR data analysed in this study was obtained from the NWO fast-track corona project (440.20.022). Requests to access this dataset should be directed to NTR data repository

## Acknowledgements

We want to thank all the study participants that have donated—and still are donating— samples to help research on COVID-19. The COVID-19 Host Genetics Initiative was originally initiated by AG and Mark Daly, but it belongs to all the participant studies.

We thank the UMCG Genomics Coordination Center, the UG Center for Information Technology and their sponsors BBMRI-NL & TarGet for storage and compute infrastructure.

We thank Kate McIntyre for editing this manuscript.

## Author contributions

Research design: IVB, PL, APSO, JMV, JCH, AG, PD, JK, HMB and LHF; Data analysis: APSO, JMV, CAW, FB, AC, MB, JJH, JK, CH, CFR, ArC, DP, ETC, KMSB, SR, AB, SW, FT, XW, JMR, YWL, JTL, NLW, EJCG, PD; Data interpretation: IVB, PL, APSO, JMV, CAW, JCH, FB, PD, LHF; IVB, PL, APSO, JMV, CAW, FB, EALM, PD wrote the manuscript with critical input from all authors. All authors read and approved the final manuscript.

## Data availability

Non-identifiable information from the GS:SFHS cohort is available to researchers in the UK and to international collaborators through application to the GS Access Committee. GS operates a managed data access process including an online application form (http://www.gsaccess.org/) and proposals are reviewed by the GS Access Committee.

The Helix data were collected under IRB Protocol #20170748. Data are available from the corresponding author on reasonable request.

The Lifelines data analysed in this study were obtained from the Lifelines biobank, under project application number ov20_0554. Requests to access this dataset should be directed to Lifelines Research Office.

The NTR data analysed in this study were obtained from the NWO fast-track corona project (440.20.022). Requests to access this dataset should be directed to NTR data repository.

## Informed consent

All biobank participants have provided informed consent that provide the opportunity for add-on research.

## Research involving human participants

Generation Scotland, Lifelines, Lifelines NEXT and Netherlands Twin Registry studies have been approved by an ethics committee. Helix data were collected under IRB Protocol #20170748.

## Lifelines COVID-19 cohort study authors

Lude H. Franke^1^, H. Marike Boezen^2^, Jochen Mierau^3,4^, Jackie Dekens^1^, Patrick Deelen^1,5^, Pauline Lanting^1^, Judith Vonk^2^, Ilja Nolte^2^, Anil Ori^1,6^, Annique Claringbould^1^, Floranne Boulogne^1^, Soesma Medema-Jankipersadsing^1^

1. University of Groningen, University Medical Center Groningen, Department of Genetics, Groningen, The Netherlands
2. University of Groningen, University Medical Center Groningen, Department of Epidemiology, Groningen, The Netherlands
3. Faculty of Economics and Business, University of Groningen, Nettelbosje 2, 9747AE Groningen
4. Aletta Jacobs School of Public Health, Groningen, the Netherlands, Landleven 1, 9747 AD Groningen, the Netherlands
5. Department of Genetics, University Medical Centre Utrecht, P.O. Box 85500, 3508 GA, Utrecht, the Netherlands
6. University of Groningen, University Medical Center Groningen, Department of Psychiatry, Groningen, The Netherlands

## The COVID-19 Host Genetics Initiative authors

### 1. BioMe

Data collection and coordination: Judy H. Cho, Ruth J.F. Loos Analysis: Arden Moscati

### 2. Corea (Genetics of COVID-related Manifestation)

Data collection and coordination: Kangbuk Samsung Cohort Study (KSCS), Yoosoo Chang, Pyoeng Gyun Choe, Jin Chung, Sinyoung Ham, Eun-Jeong Joo, Jongtak Jung, Chang Kyung Kang, Hyung-Lae Kim, Hong Bin Kim, Eu Suk Kim, Hyo-Jung Lee, Soo-kyung Park, Kyoung-Un Park, Jeong Su Park, Seungho Ryu, Kyoung-Ho Song

Technical support: Global Science Experimental Data Hub Center (GSDC), Korea Research Environment Open Network (KREONET)

Analysis: Han-Na Kim

Administrative support: Nam-Jong Paik

### 3. COVID19-Host(a)ge

Ethics and communication: Agustín Albillos, Rosanna Asselta, Luis Bujanda, Maria Buti, Stefano Duga, Javier Fernández, Manuel Romero Gomez, Pietro Invernizzi, Daniele Prati

Data collection and coordination: Jesus M. Banales, Trine Folseraas, Andre Franke, Johannes R Hov, Tom H Karlsen, Luca Valenti

Analysis: Frauke Degenhardt, David Ellighaus

### 4. deCODE genetics

Data collection and coordination: Elias S Eythorsson, Asgeir Haraldsson, Dadi Helgason, Hilma Holm, Ragnar F Ingvarsson, Ingileif Jonsdottir, Gudmundur L Norddahl, Runolfur Palsson, Jona Saemundsdottir, Kari Stefansson, Unnur Thorsteinsdottir

Analysis: Daníel F Gudbjartsson, Hakon Jonsson, Pall Melsted, Patrick Sulem, Gardar Sveinbjornsson

### 5. Determining the Molecular Pathways and Genetic Predisposition of the Acute Inflammatory Process Caused by SARS-CoV-2

Data collection and coordination: Marta E. Alarcón-Riquelme, David Bernardo, Silvia Rojo Rello Analysis: Manuel Martínez-Bueno

### 6. Finngen

Data collection and coordination: Finngen

### 7. GEN-COVID, reCOVID

Technical support: Sara Amtrano, Mirella Bruttini, Valentino Floriana, Anna Rita Giliberti Analysis: Elisa Benetti, Chiara Fallerini, Simone Furini, Anna Maria Pinto

Data collection and coordination: Margherita Baldassarri, Francesca Fava, Francesca Mari, Alessandra Renieri

Other: Susanna Croci, Rossella Tita

Ethics and communication: Elisa Frullanti

### 8. Generation Scotland

Analysis: Drew Altshul, Archie Campbell, Caroline Hayward

Data collection and coordination: Chloe Fawns-Ritchie, David Porteous

### 9. Genes & Health

Data collection and coordination: Qinqin Huang, Karen A Hunt, Hilary C Martin, Dan Mason, Richard C Trembath, Bhavi Trivedi, John Wright

Other: Sarah Finer, Christopher Griffiths

Analysis: David A van Heel

### 10. Genetic determinants of COVID-19 complications in the Brazilian population

Data collection and coordination: Cinthia E Jannes, Jose E Krieger, Alexandre C Pereira Technical support: Emmanuelle Marques

### 11. Genomic epidemiology of SARS-Cov-2 and host genetics in Coronavirus Disease 2019 (COVID-19)

Technical support: Karen Dalton, Christopher DeBoever, Illumina, Inc., David Jimenez-Morales, Aldo Cordova Palomera, Benjamin Pinsky, Erin Smith, Sandor Szalma, Cathy Tralau-Stewart, Emily Wong

Data collection and coordination: John Gorzynski, Hannah de Jong

Analysis: David Amar, Olivier Delaneau, Christopher Hughes, Alexander Ioannidis, Archana Raja, Simone Rubinacci, Yosuke Tanigawa

Administrative support: Euan Ashley, Carlos Bustamante, Vicki Parikh, Manuel Rivas, Matthew Wheeler

### 12. Genetic modifiers for COVID-19 related illness

Data collection and coordination: Adeline Busson, Jean-Christophe Goffard, Isabelle Migeotte, Xavier Peyrassol, Guillaume Smits, Isabelle Vandernoot, Francoise Wilkin

Technical support: Youssef Bouysran, Bruno Pichon, Nicky Tiembe

### 13. Helix Exome+ COVID-19 Phenotypes

Data collection and coordination: Kelly M. Schiabor Barrett, Alexandre Bolze, Elizabeth T. Cirulli, Jimmy M. Ramirez III, Yan Wei Lim, James T. Lu, Stephen Riffle, Francisco Tanudjaja, Xueqing Wang, Nicole L. Washington, Simon White

### 14. Lifelines

Analysis: Annique Claringbould, Patrick Deelen, Esteban Lopera, Robert Warmerdam Data collection and coordination: Marike Boezen, Lude Franke

### 15. Mass General Brigham – Host Vulnerability to COVID-19

Data collection and coordination: Robert Green, Beth Karlson, James Meigs, Josep Mercader, Shawn Murphy, Emma Perez, Sue Slaugenhaupt, Jordan Smoller, Scott Weiss, Ann Woolley Analysis: Yen-Chen Anne Feng

### 16. Netherlands Twin Register

Data collection and coordination: Meike Bartels, Eco de Geus, Michel G Nivard Analysis: Jouke-Jan Hottenga

### 17. Population controls

Data collection and coordination: Alfredo Brusco, Cynthia M Bulik, Denis Franchimont, Mikael Landen, Edouard Louis, Nancy Pedersen, Souad Rahmouni, Pasquale Striano, Severine Vermeire, Federico Zara

### 18. Qatar Genome Program

Data collection and coordination: Wadha Al-Muftah, Radja Badji, Said Ismail Analysis: Yasser Al-Sarraj, Hamdi Mbarek

### 19. SIGMA

Data collection and coordination: Marta E. Alarcón-Riquelme

### 20. UK 100,000 Genomes Project

Data collection and coordination: Prabhu Arumugam, Mark Caulfield, Genomics England Research Consortium, Anna C Need, Thomas Oscroft, Augusto Rendon, Richard H Scott Analysis: Georgia Chan, Athanasios Kousathanas, Loukas Moutsianas, Chris A Odhams, Dorota Pasko, Dan Rhodes, Alex Stuckey

### 21. UK Biobank

Analysis: Elizabeth G. Atkinson, Nikolas Baya, Guillaume Butler-Laporte, Hilary Finucane, Vincenzo Forgetta, Masahiro Kanai, Konrad J. Karczewski, Nils Koelling, Alicia R. Martin, Tomoko Nakanishi, Duncan S. Palmer, J. Brent Richards, Chris C A Spencer, Patrick Turley, Raymond K. Walters, Daniel J Wilson

Data collection and coordination: Jacob Armstrong, Anne Marie O’Connell, David H Wyllie

Technical support: Sam Bryant

Administrative support: Claire Churchhouse

### 22. UK Blood Donors Cohort

Data collection and coordination: Emanuele Di Angelantonio, Michael Chapman, John Danesh, Willem Ouwehand, Dave Roberts, Nick Watkins

Analysis: Adam Butterworth, Jing Hua Zhao

### 23. COVID-19 Host Genetics Initiative Coordination

Phenotype steering group: Les Biesecker, Lea Davis, Patrick Deelen, Andrea Ganna, David van Heel, Eric Kerchberger, Sulggi Lee, Tomoko Nakanishi, James Priest, Alessandra Renieri, Brent Richards, Vijay Sankaran

Administrative support: Karolina Chwialkowska, Margherita Francescatto, Christine Stevens International Common Disease Alliance: Amy Trankiem, Kate Balaconis

Leadership: Rachel Liao, Mark Daly, Andrea Ganna, Ben Neale

Data dictionary: Anna Bernasconi, Stefano Ceri, Francesca Mari, Alessandra Renieri

Analysis: Juha Karjalainen, Mattia Cordioli, Mari Niemi, Wei Zhou

Website: Huy Nguyen, Matthew Solomonson

### 24. In silico follow-up results

Analysis: Hilary Finucane, Shyamalika Gopalan, Kangcheng Hou, Philip Jansen, Masahiro Kanai, Christiaan de Leeuw, Zeyun Lu, Nicholas Mancuso, Eirini Marouli, Areti Papadopoulou, Bogdan Pasaniuc, Gita Pathak, Renato Polimanti, Danielle Posthuma, Jeanne Savage, Emil Uffelmann, Peter Visscher, Frank R Wendt, Naomi Wray, Loic Yengo

## Supplementary information

### Supplementary Methods

Detailed information on cohort level GWAS and C19HG meta-analysis.

### Supplementary Figures

**Figure S1.** Chronic disease associations with COVID-19 predicted cases and COVID-19 positive cases. Fisher’s exact test shows a Bonferroni significant positive correlation between “Neurological disease”, “Psychological disease”, “Chronic muscle disease”, “Cancer” and “Lung disease” patients and COVID-19 predicted cases. This association is not present for positive COVID-19 cases. The same diseases are Bonferroni significant and not significant, respectively, when applying generalised linear models with “age”, “sex” and “bmi” as confounding variables (not shown).

**Figure S2.** Regional association plots for the locus of the two top SNPs rs11844522 (A) and rs5798227 (B) in the predicted COVID-19 GWAS (≤ 1×10^−6^). In each of the two panels, the top SNP is indicated by a purple diamond. Other SNPs are colored according to their linkage disequilibrium with the top SNP (calculated with the European population from the 1000 Genomes Project (phase 3) as a reference panel). The genes located within the visualized regions are drawn at their respective locations, with an arrow indicating the transcribed strand. Positions correspond to genome assembly GRCh37.

**Figure S3.** Comparison of genome-wide significant independent SNPs discovered in other viral infection phenotypes replicated in the four COVID-19 phenotypes *B2*: Hospitalized COVID-19 vs. population, *C1*: COVID-19 vs. self-reported negative, *C2*: COVID-19 vs. population, *D1*: Predicted COVID-19 from self-reported symptoms vs. predicted or self-reported non-COVID-19.

**Figure S4.** Quantile-quantile plots (Q-Q plots) of the four phenotypes: *B2*: Hospitalized COVID-19 vs. population, *C1*: COVID-19 vs. self-reported negative, *C2*: COVID-19 vs. population and *D1*: Predicted COVID-19 from self-reported symptoms vs. predicted or self-reported non-COVID-19, wherein variants are confined to a selection of associated with several viral infection traits. For every COVID-19 GWAS, a genomic inflation factor (λ) with accompanying pvalue is shown. Based on both these values and the Q-Q plots, there is no indication of an increased viral infection signal in one of the COVID-19 phenotypes.

### Supplementary Tables

**Table S1.** Symptoms with respective cut-off values used in the Generation Scotland, Helix, Lifelines and NTR cohorts. Logistic regression was performed for each symptom separately in Lifelines on positive (n = 56) vs negative (n = 586) tested individuals to define symptom cut-offs. The odds ratios and p-values of used cut-offs in Lifelines are displayed here.

**Table S2.** The different cut-offs and symptoms that were used by the Generation Scotland, Helix, Lifelines and NTR cohorts when applying the Menni COVID-19 prediction model to the datasets prior to running the GWAS.

**Table S3.** Full phenotype description and detailed analysis plan used within the C19HG for genome-wide association analysis

**Table S4.** Descriptive statistics of the Helix, Lifelines and NTR cohorts. Due to absence of testing data, Generation Scotland could not be used for replication of the Menni COVID-19 prediction model and in the development of the Lifelines COVID-19 prediction model.

**Table S5.** Reported number of individuals with an essential occupation and who have been in contact with infected individuals across negatively and positively tested COVID-19 cases in the Lifelines cohort.

**Table S6.** Overlapping symptoms in the Helix, Lifelines and NTR cohorts. Logistic regression for each symptom separately in Lifelines on positive (n = 56) vs negative (n = 586) tested subjects to define symptom cut-offs (reference = absence of symptom).

**Table S7a.** Overlapping symptoms in the Helix, Lifelines and NTR cohorts. Logistic regression for each symptom separately in Lifelines on positive (n = 56) vs negative (n = 586) tested subjects to define symptom cut-offs (reference = absence of symptom).

**Table S7a**. The Lifelines COVID-19 prediction model.

**Table S7b**. Diagnostics of different cut-offs of predicted probability of the Lifelines COVID-19 prediction model.

**Table S7c**. Model diagnostics of the Lifelines COVID-19 prediction model in the Helix, Lifelines and NTR cohorts.

**Table S8.** Comparison of the case predictions of the Lifelines COVID-19 prediction model and the Menni COVID-19 prediction model in the Helix, Lifelines and NTR cohorts.

**Table S9**. Downstream analysis of DEPICT

## References

1. World Health Organization. WHO Coronavirus Disease (COVID-19) Dashboard. https://covid19.who.int/.

2. World Health Organization. Coronavirus – Symptoms. https://www.who.int/health-topics/coronavirus#tab=tab_3.

3. Vabret, N. et al. Immunology of COVID-19: Current State of the Science. Immunity 52, 910–941 (2020).

4. Gupta, A. et al. Extrapulmonary manifestations of COVID-19. Nat. Med. 26, 1017–1032 (2020).

5. Rubicz, R. et al. Genetic Factors Influence Serological Measures of Common Infections. Hum. Hered. 72, 133–141 (2011).

6. Grundbacher, J. Heritability Estimates and Genetic and Environmental Correlations for the Human Immunoglobulins G, M, and A. 12.

7. Tian, C. et al. Genome-wide association and HLA region fine-mapping studies identify susceptibility loci for multiple common infections. Nat. Commun. 8, 599 (2017).

8. The COVID-19 Host Genetics Initiative. The COVID-19 Host Genetics Initiative, a global initiative to elucidate the role of host genetic factors in susceptibility and severity of the SARS-CoV-2 virus pandemic. Eur. J. Hum. Genet. 28, 715–718 (2020).

9. Menni, C. et al. Real-time tracking of self-reported symptoms to predict potential COVID-19. Nat. Med. 26, 1037–1040 (2020).

10. Pers, T. H. et al. Biological interpretation of genome-wide association studies using predicted gene functions. Nat. Commun. 6, 5890 (2015).

11. Cai, N. et al. Minimal phenotyping yields genome-wide association signals of low specificity for major depression. Nat. Genet. 52, 437–447 (2020).

12. Wen, W. et al. Immune cell profiling of COVID-19 patients in the recovery stageby single-cell sequencing. Cell Discov. 6, 31 (2020).

13. Lu, J. & Bowes, J. Introducing the Helix DNA Discovery Project. https://blog.helix.com/helix-dna-discovery-project/#:~:text=April%205%2C%202018-,Introducing%20the%20Helix%20DNA%20Discovery%20Project,improve%20their%20life%20through%20DNA.&text=The%20project%20is%20entirely%20voluntary,used%20to%20study%20human%20genetics.

14. C19HG. COVID-19 hostgene initiative questionnaires. https://docs.google.com/document/d/12O6h5EcVCb7y3w8vJPEef1Tpjg0x2fmge9c1uhYTlfo/edit (2020).

15. Mc Intyre, K. et al. The Lifelines COVID-19 Cohort: a questionnaire-based study to investigate COVID-19 infection and its health and societal impacts in a Dutch population-based cohort. http://medrxiv.org/lookup/doi/10.1101/2020.06.19.20135426 (2020) doi:10.1101/2020.06.19.20135426.

16. Helix. Variants Pipeline Performance White Paper. https://cdn.shopify.com/s/files/1/2718/3202/files/Helix_Performance_White_Paper_v4.pdf (2019).

17. Kendig, K. I. et al. Computational performance and accuracy of Sentieon DNASeq variant calling workflow. http://biorxiv.org/lookup/doi/10.1101/396325 (2018) doi:10.1101/396325.

18. Cirulli, E. T. et al. Genome-wide rare variant analysis for thousands of phenotypes in over 70,000 exomes from two cohorts. Nat. Commun. 11, 542 (2020).

19. The 1000 Genomes Project Consortium. A global reference for human genetic variation. Nature 526, 68–74 (2015).

20. Zheng, X. & Davis, J. SAIGEgds – an efficient statistical tool for large-scale PheWAS with mixed models. (2019).

21. Zhou, W. et al. Efficiently controlling for case-control imbalance and sample relatedness in large-scale genetic association studies. Nat. Genet. 50, 1335–1341 (2018).

22. Buniello, A. et al. The NHGRI-EBI GWAS Catalog of published genome-wide association studies, targeted arrays and summary statistics 2019. NHGRI-EBI GWAS Catalog https://www.ebi.ac.uk/gwas/docs/about (2019).

23. Sanna, S. et al. UGLI (release 1.0) Quality Control Report. https://drive.google.com/file/d/1qzD6ZOqyTjYMH89dM051L3G0qTVQ0QYP/view.

